# Racial Differences in Temporal Thermometry and Association with Delayed Sepsis Bundle Care

**DOI:** 10.1101/2025.03.31.25324893

**Authors:** João Matos, Mahmoud Alwakeel, Sicheng Hao, Inês Martins, Jaime S. Cardoso, Judy Wawira Gichoya, Leo Anthony Celi, Andrea Lane, Vijay Krishnamoorthy, Sivasubramanium V. Bhavani, Christopher E. Cox, Warren A. Kibbe, Chuan Hong, A.Ian Wong

## Abstract

**Importance:** Early identification of fever or hypothermia is crucial for diagnosing sepsis. Despite their increased use across healthcare systems, concerns have been raised about the accuracy of temporal thermometers among Black patients.

**Objective:** To study the performance of temporal thermometry across race and ethnicity, and its impact on the initiation of the sepsis management bundle (SEP-1).

**Design:** In this retrospective cohort study, records from 2008–2024 in 123 U.S. hospitals were analyzed, including electronic health records from Duke, MIMIC-IV, eICU-CRD-1, and eICU-CRD-2. Patients were included if they had a temporal measurement within one hour of an oral/core measurement (esophageal, bladder, rectal).

**Main Outcomes and Measures:** *Hidden Fever* was defined when the temporal thermometer read ≤ 38°C but oral/core measurement > 38°C and *Hidden Hypothermia* as temporal temperature ≥ 36°C but oral/core < 36°C. The primary outcome was *Hidden Hyper/Hypothermia (HHH)* when either *Hidden Fever* or *Hidden Hypothermia* happened. *Observed Hyper*/*Hypothermia (OHH)* corresponded to an agreement between both measurements, used as a control. To study the impact of temporal thermometry underperformance on SEP-1 bundle initiation, we assessed time to culture ordering, antibiotics administration, and lactate measurement. A composite of these three, whichever happened first, was defined as our secondary outcome.

**Results:** 6,921 paired temperature measurements were studied for the primary outcome analysis, corresponding to 4,248 hospitalizations (2.6% Asian; 12.4% Black; 2.6% Hispanic/Latino; 83.4% White; 44.9% female; 7.7% with HHH) from 115 hospitals. After adjusting for confounders, Black [OR (95% CI): 1.760 (1.219, 2.541), *p*=0.003] and Hispanic/Latino [OR (95% CI): 2.183 (1.226, 3.888), *p=*0.008] patients were significantly more likely to present with HHH than White patients.

For the study of the secondary outcome, 434 patients had either OHH or HHH. Compared to patients with OHH, patients with HHH had significantly delayed secondary outcomes, with log-rank p-value of 0.002.

**Conclusions and Relevance:** Solely relying on temporal thermometry can lead to missed hyper/hypothermia events, especially among Asian, Black, and Hispanic patients. This has been associated with delays in the initiation of the SEP-1 bundle, which may hamper health systems’ compliance with reimbursement programs.

**Key Points:** *Question:* Are there racial differences in temporal thermometry that result in missed hyper/hypothermia, and are these associated with delays in sepsis bundle care (SEP-1) initiation?

*Findings:* In this multicenter retrospective cohort study of 4,248 hospitalizations, Asian, Black, and Hispanic patients were more likely to have hidden hyper/hypothermia when using temporal thermometers, as compared to oral/core measurements. These missed events were associated with delayed SEP-1 interventions (three out of four were studied: culture orders, antibiotic administration, and lactate measurements).

*Meaning:* Reliance on temporal thermometry could exacerbate health disparities and hamper hospital systems’ compliance with reimbursement programs. Temporal thermometers should therefore be routinely validated with contact thermometers.

## Background

Since their introduction into the medical field 25 years ago, use of temporal thermometers has substantially increased across healthcare systems due to their low cost and ease of use.^1^ Consequently, they became the most commonly used method for screening billions of people during the COVID-19 pandemic.^2^ For healthcare providers, early identification of fever (body temperature > 38.0°C) or hypothermia (body temperature < 36.0°C) using strict temperature cutoffs are crucial for recognizing patients who may meet the systemic Inflammatory Response Syndrome (SIRS) temperature criteria, which can contribute to identifying those at risk for sepsis in conjunction with other clinical findings. According to the Centers for Medicare & Medicaid Services (CMS), sepsis is diagnosed by the presence of a suspected or confirmed infection alongside at least two SIRS criteria, which include elevated or lowered temperature, rapid heart rate, increased respiratory rate, and abnormal white blood cell count. These established cut-off temperatures are strict and important for timely sepsis care; for example, electronic notification systems often trigger notifications and management bundles if the temperature at 38.1°C but not at 37.9°C, concomitantly with another positive SIRS criteria.^3^ Promptly identifying and initiating the severe sepsis and septic shock management bundle (SEP-1) — which includes blood cultures, antibiotics, fluids, and lactate measurement — within the first hour is associated with lower hospital mortality rates and better outcomes.^4–6^ In 2024, the Centers for CMS incorporated SEP-1 compliance into the Hospital Value-Based Purchasing (VBP) program. Under this program, early sepsis identification and adherence to SEP-1 guidelines are critical to ensure hospitals do not lose reimbursement funds.^7^

While the gold standard for temperature measurement is to measure core body temperature using invasive probes located in the pulmonary artery, esophagus, bladder, or rectum, these methods are not routinely used in daily practice due to their cost, inconvenience, resource requirements, and increased risk of infection and complications.^8^ Therefore, guidelines and hospitals continue to use oral temperature measurements as the reference for temperature threshold recommendations, ensuring standardization with one of the most commonly available and convenient methods.^9^

The process of measuring temporal temperature involves scanning the skin using infrared light and subsequently compensating for heat loss to the surrounding environment.^10^ Due to the use of infrared light, similar to pulse oximetry, recent concerns have emerged regarding potential device bias against minority patient populations.^11^ Bhavani et al.^11^, found that temporal artery thermometer readings were lower than oral temperature readings in Black patients but not in White patients, impacting fever identification rates among over 4,000 hospitalized patients at Emory University. These concerns are becoming more critical due to the growing evidence that pulse oximetry has been implicated in contributing to racial disparities in healthcare due to inaccurate measurements, leading to delays in disease recognition and management, as well as increased end-organ damage and mortality.^12–15^ Asian, Black, and Hispanic patient populations constitute one-third of severe sepsis and septic shock patients.^16^ Any delays in identifying fever or hypothermia as signs of sepsis would compromise SEP-1 compliance and affect reimbursement under the VBP program.

While the Bhavani et al.^11^ study is the first in the literature to highlight the potential racial disparities associated with temporal temperature measurement, the study was restricted to Black and White populations from a single health system, did not examine the potential disparity in detecting hypothermia (just fever), and did not investigate the potential association of this disparity with standardized patient outcomes. Therefore, our study builds upon the prior study by examining differences in temporal thermometry readings versus oral/core temperature readings among hospitalized patients of different races and ethnicities. Additionally, we evaluate the clinical implications of these disparities on the potential early initiation of the SEP-1 bundle.

## Methods

This study follows the American Medical Association’s recommendations on health equity language and adheres to the STROBE statement.^17,18^ Analyses were performed using R 4.2, Python 3.8, and SQL.

### Data Sources

Four electronic health records (EHR) databases were studied: Duke Health (2014-2024), MIMIC-IV (2008-2022),^19^ eICU-CRD-1 (2014-2015),^20^ and eICU-CRD-2 (2019-2022). This study was approved by the Duke Health IRB under Pro00110102. The latter three datasets have been previously de-identified and did not require a waiver for informed consent. (Supplemental text)

### Paired Temperature Measurements

Temperature pairs are defined when a temporal measurement occurs one hour before an oral/core temperature measurement (esophageal, bladder, rectal).

### Outcomes

The primary outcome was Hidden Hyper/Hypothermia (HHH), which can be defined as either Hidden Hyperthermia or Hidden Hypothermia. Hidden Hypothermia occurs when oral/core measurement > 38.0°C, but the temporal measurement ≤ 38.0°C; Hidden Hypothermia when the oral/core measurement < 36.0°C, but the temporal measurement ≥ 36.0°C. These can also be interpreted as false negatives. As control outcomes, Observed Hyper/Hypothermia (OHH) corresponded to an agreement between temporal and oral/core measurements in detecting hyperthermia or hypothermia, i.e., true positives. (Supplemental eFigure 3)

The secondary outcome was time to culture ordering, antibiotic administration, or lactate measurement ordering (whichever occurred first, covering three of the four SEP-1 bundle actions). Time zero was defined as the time of temporal temperature measurement. We assumed that patients with HHH met the CMS definition if hyperthermia or hypothermia were identified, even though this may not apply to all cases. This assumption was adopted for exploratory purposes, as extracting all remaining SIRS criteria and determining whether the patient has a suspected or confirmed infection is challenging.

### Hypothesis

The primary hypothesis was that HHH’s prevalence varies per race and ethnicity (a surrogate for skin tone). The secondary hypothesis was that patients with HHH had a delayed initiation of any of the three aforementioned SEP-1 bundle actions.

### Cohort Selection

Adult patients were included if at least one paired temperature measurement (temporal reading followed by oral/core measurement) was present at any time during their hospitalization.

For the primary outcome (HHH), while maintaining a homogeneous cohort (mostly ICU), patients without a first day SOFA score were excluded. Additionally, patients whose self-reported race and ethnicity did not fall into “Asian”, “Black”, “Hispanic/Latino”, or “White” were excluded.

For the secondary outcome (SEP-1 initiation), only first pairs from patients with HHH *or* OHH were included, and hospitals without microbiology, medication, or lactate measurement data (eICU-CRD) were excluded.

### Missing Data Handling

Patients who had missing data for any of the covariates mentioned above (thus preventing us from applying the inclusion and exclusion criteria) were not included in the study. Missing data on treatment initiation was assumed to imply absence of treatment initiation.

### Statistical analysis

As an exploration analysis, temporal and oral/core measurements’ distributions were compared between White (reference) and “Asian”, “Black”, and “Hispanic/Latino” groups, across different reference temperature thresholds, separately, using t-tests.

To determine the likelihood of *HHH* across race and ethnicity, generalized estimating equations were fit, with an “independence” correlation structure, adjusting for age, sex, first-day SOFA score, and database. Hospital admission identifiers were used as a grouping factor to account for potential correlations among pairs captured within the same hospitalization. Differences were reported in terms of adjusted odds ratios (OR). Null hypotheses were rejected at *p* < 0.05. Sex was additionally considered as a negative control disparity axis.

To determine differences in time to SEP-1 bundle onset (3 out of 4 actions, due to data limitations) among patients with *HHH*, compared to *OHH*, cumulative incidence curves were plotted using a Kaplan-Meier estimator. A log-rank test was performed to compare both curves (HHH patients versus OHH). For each model, the outcome of interest was the time to the presence of each of the aforementioned SEP-1 interventions. Observations were right-censored upon ICU discharge or 24 hours after the temperature pair, whichever came first. Null hypotheses were rejected at *p* < 0.05. Finally, culture positivity and measured lactate levels were compared between the study and control groups.

## Results

### Cohort Characteristics – primary outcome

6,921 paired temperature measurements were identified in total, corresponding to 4,144 patients, 4,248 hospitalizations. (Figure 1) Of these hospitalizations, 59 (1.4%) were from Duke Health; 884 (20.8%) from MIMIC-IV; 1,793 (42.2%) from eICU-CRD-1; and 1,512 (35.6%) from eICU-CRD-2. In this cohort, 111 (2.6%) of the patients were Asian; 483 (11.4%) Black; 112 (2.6%) Hispanic/Latino; 3,543 (83.4%) White; and 1,907 (44.9%) Female. 325 (7.7%) hospitalizations had at least one temperature pair with HHH. (Table 1) The median [IQR] SOFA was 5.0 [3.0,8.0], and similar across race and ethnicity.

**Figure 1.**
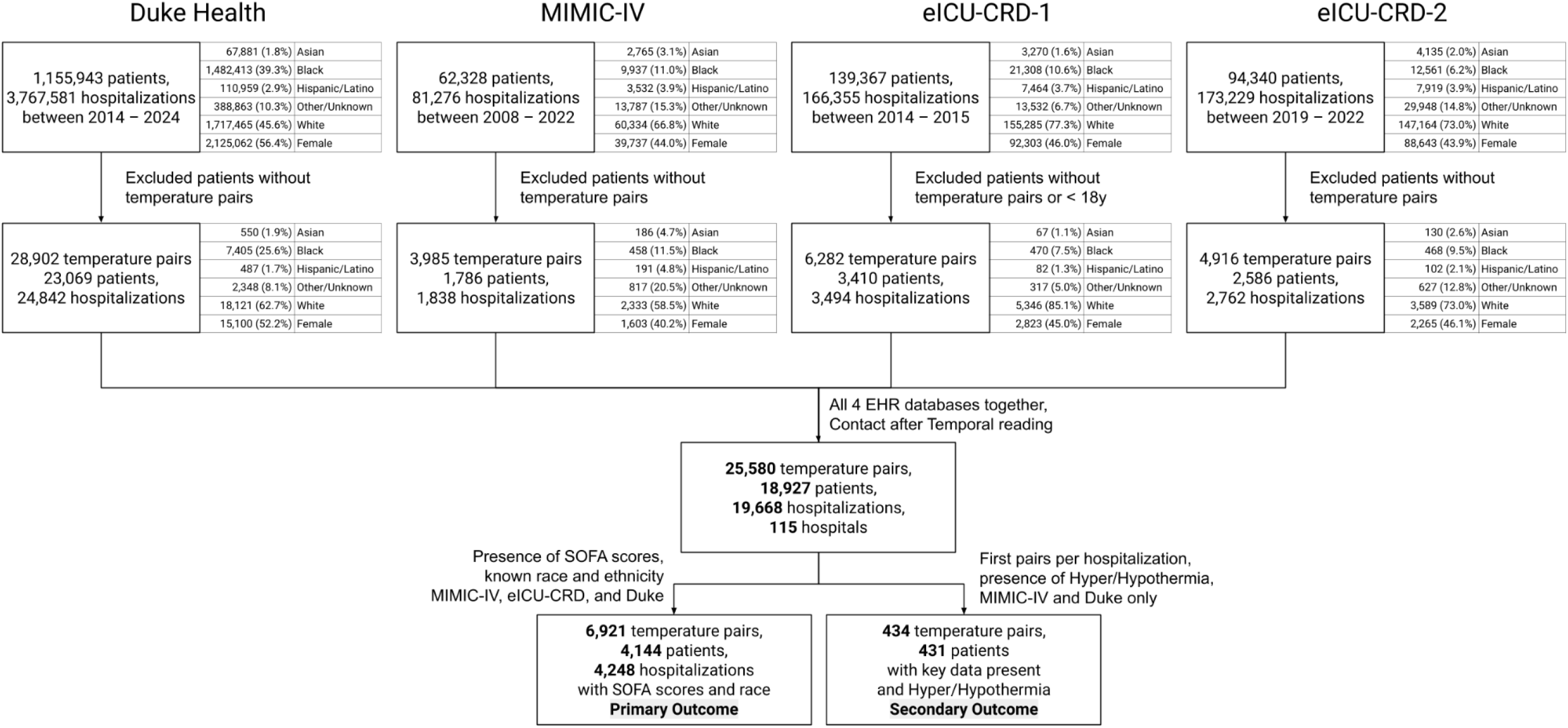
Cohort Selection Flow Diagrams. After applying our exclusion criteria, we obtained 4,248 hospitalizations for the primary outcome analysis and 431 patients/hospitalizations for the secondary outcome analysis. At Duke Health, a drop from 39.3% to 25.6% was verified in the Black subpopulation, when patients without a temperature pair were excluded from the study, potentially indicating an underrepresentation of these patients as the exclusion criteria were applied.

**Table 1.** Cohort characteristics (primary outcome), grouped by race and ethnicity, per hospitalization.

|  | Grouped by Race and Ethnicity |  |  |  |  |  |
| --- | --- | --- | --- | --- | --- | --- |
|  | Missing | Overall | Asian | Black | Hispanic/Latino | White |
| N (%) |  | 4,248 | 111 (2.6) | 483 (11.4) | 112 (2.6) | 3,542 (83.4) |
| Age (admission, years), median [Q1,Q3] | 0 | 64.0 [52.0,74.0] | 63.0 [52.0,71.5] | 62.0 [50.0,72.0] | 58.5 [51.8,68.0] | 65.0 [52.2,74.0] |
| SOFA, day 1 (24h), median [Q1,Q3] | 0 | 5.0 [3.0,8.0] | 5.0 [2.5,9.0] | 6.0 [4.0,8.5] | 5.0 [3.0,9.0] | 5.0 [3.0,8.0] |
| ICU LoS (days), median [Q1,Q3] | 1 | 2.9 [1.8,6.6] | 4.4 [2.0,10.9] | 3.7 [2.0,10.0] | 4.3 [2.7,10.3] | 2.8 [1.7,5.9] |
| Hospital LoS (days), median [Q1,Q3] | 0 | 10.1 [5.3,19.9] | 12.7 [6.0,29.4] | 13.8 [7.0,27.4] | 12.0 [7.5,22.2] | 9.8 [5.1,18.2] |
| Female Sex, n (%) | 0 | 1907 (44.9) | 54 (48.6) | 233 (48.2) | 51 (45.5) | 1569 (44.3) |
| Database, n (%) |  |  |  |  |  |  |
| Duke | 0 | 59 (1.4) | 2 (1.8) | 30 (6.2) |  | 27 (0.8) |
| MIMIC-IV+ |  | 884 (20.8) | 45 (40.5) | 126 (26.1) | 50 (44.6) | 663 (18.7) |
| eICU-1 |  | 1793 (42.2) | 17 (15.3) | 156 (32.3) | 26 (23.2) | 1594 (45.0) |
| eICU-2 |  | 1512 (35.6) | 47 (42.3) | 171 (35.4) | 36 (32.1) | 1258 (35.5) |
| Year, n (%) |  |  |  |  |  |  |
| 2008 - 2010 | 0 | 89 (2.1) | 2 (1.8) | 27 (5.6) | 7 (6.2) | 53 (1.5) |
| 2011 - 2013 |  | 85 (2.0) | 1 (0.9) | 11 (2.3) | 8 (7.1) | 65 (1.8) |
| 2014 - 2016 |  | 1967 (46.3) | 35 (31.5) | 171 (35.4) | 30 (26.8) | 1731 (48.9) |
| 2017 - 2019 |  | 243 (5.7) | 8 (7.2) | 24 (5.0) | 14 (12.5) | 197 (5.6) |
| 2020 - 2022 |  | 1849 (43.5) | 64 (57.7) | 240 (49.7) | 53 (47.3) | 1492 (42.1) |
| 2023 - 2024 | 15 (0.4) | 1 (0.9) | 10 (2.1) |  | 4 (0.1) |  |
| At least one pair in the ICU, n (%) | 0 | 2895 (68.1) | 86 (77.5) | 373 (77.2) | 92 (82.1) | 2344 (66.2) |
| Hidden Fever (at least once), n (%) | 0 | 169 (4.0) | 6 (5.4) | 28 (5.8) | 9 (8.0) | 126 (3.6) |
| Observed Fever (at least once), n (%) | 0 | 151 (3.6) | 3 (2.7) | 23 (4.8) | 8 (7.1) | 117 (3.3) |
| Hidden Hypothermia (at least once), n (%) | 0 | 157 (3.7) | 7 (6.3) | 25 (5.2) | 8 (7.1) | 117 (3.3) |
| Observed Hypothermia (at least once), n (%) | 0 | 174 (4.1) | 5 (4.5) | 39 (8.1) | 9 (8.0) | 121 (3.4) |
| HHH (at least once), n (%) | 0 | 325 (7.7) | 13 (11.7) | 53 (11.0) | 17 (15.2) | 242 (6.8) |
| OHH (at least once), n (%) | 0 | 324 (7.6) | 7 (6.3) | 62 (12.8) | 17 (15.2) | 238 (6.7) |
| In-Hospital Mortality, n (%) | 7 | 518 (12.2) | 16 (14.4) | 78 (16.1) | 24 (21.6) | 400 (11.3) |

Summary characteristics of the first cohort, stratified by race and ethnicity, to study the primary outcome: HHH.

### Cohort characteristics – secondary outcome

434 patients (one temperature pair per patient) were found to have HHH (236, 54.5%) or OHH (198, 45.6%) in their first temperature pair, in hospitals where microbiology, medication, and lactate measurement data were available. Overall SOFA score at the time of the temperature pair was 6.0 [4.0,10.0]. For these patients, overall and after the pair and before hospital discharge, 110 (25.3%) patients had cultures ordered, 258 (59.4%) antibiotics administered, and 142 (32.7%) lactate measured. (Table 3)

### Exploratory comparison of Temporal versus Oral/Core Measurements

In critical thresholds (temporal readings close to 36.0 or 38.0°C), the oral/core measurements are found to be more out of the 36–38°C range for Asian, Black, and Hispanic/Latino patients, when compared to White patients. (Supplemental eFigure 1) To illustrate this, in the (37.5, 38.0] °C temporal temperature bin, the median [Q1,Q3] oral/core temperature reading is 37.3 [36.9, 37.8] °C for White patients, but this appears elevated for Black patients, at 37.5 [37.1, 38.0] °C (*p* < 0.001, comparing Black versus White patients’ oral/core temperature readings), despite no differences in the temporal thermometry distributions, with a median [Q1, Q3] of 37.7 [37.6, 38.8] °C for both White and Black patients (*p* = 0.803, comparing Black and White patients’ temporal temperature readings). A similar analysis using the root mean squared error (RMSE), in Supplemental eFigure 2 suggests similar findings.

**Figure 2.**
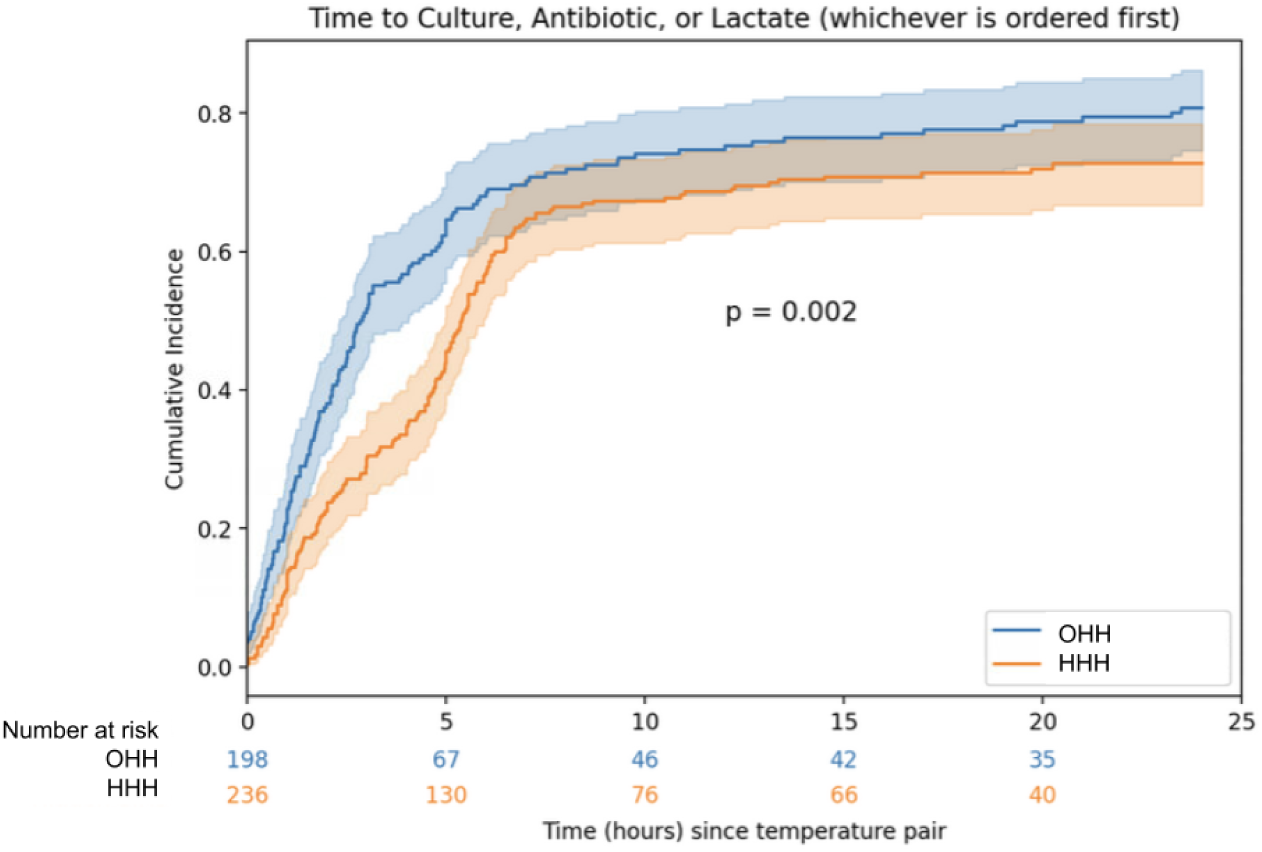
Time to culture, antibiotic, or lactate (whichever is ordered first). Limited to first pairs and temporal reading first. Unadjusted time-to-event analysis. The outcome is defined as a composite of culture order, or antibiotic administration, or lactate measurement, whichever happens first, before hospital discharge. The cumulative incidence curves were plotted using a Kaplan-Meier estimator. A log-rank test was performed to compare both curves (HHH patients versus OHH).

### Primary outcome: Hidden Hyper/Hypothermia (HHH)

After adjusting for confounders, Black [OR (95% CI): 1.760 (1.219, 2.541), *p*=0.003] and Hispanic or Latino [OR (95% CI): 2.183 (1.226, 3.888), *p=*0.008] patients showed higher odds of experiencing HHH phenomena than White patients. The directionality of these findings is consistent for Asian patients as well, although not significant. Furthermore, there were no significant differences in HHH between female and male patients, OR (95% CI): 1.149 (0.869, 1.518), *p*=0.33. (Table 2)

**Table 2.**
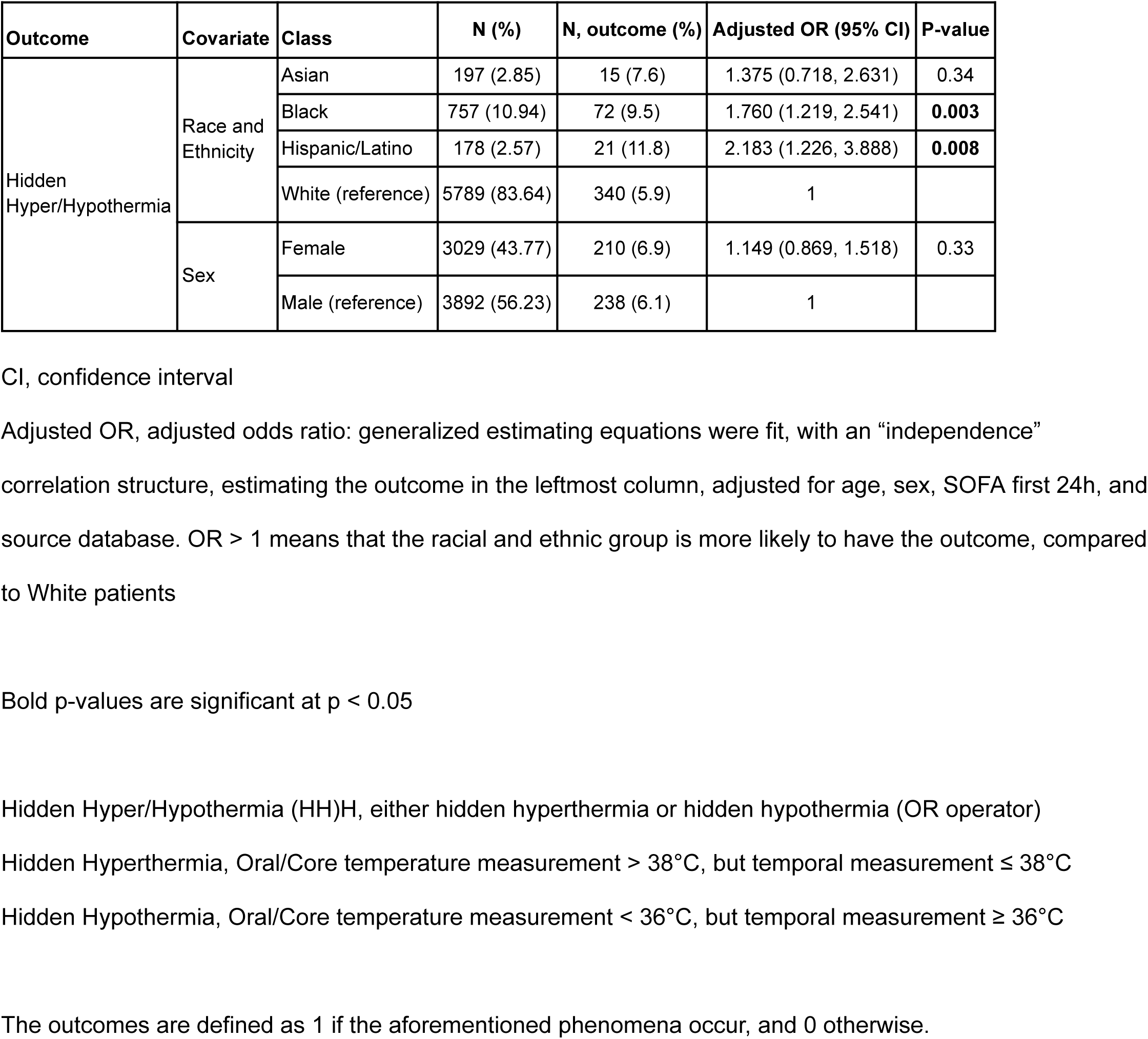
Adjusted odds ratios of HHH, across race and ethnicity and sex groups.

| Outcome | Covariate | Class | N (%) | N, outcome (%) | Adjusted OR (95% CI) | P-value |
| --- | --- | --- | --- | --- | --- | --- |
| Hidden Hyper/Hypothermia | Race and Ethnicity | Asian | 197 (2.85) | 15 (7.6) | 1.375 (0.718, 2.631) | 0.34 |
|  |  | Black | 757 (10.94) | 72 (9.5) | 1.760 (1.219, 2.541) | **0.003** |
|  |  | Hispanic/Latino | 178 (2.57) | 21 (11.8) | 2.183 (1.226, 3.888) | **0.008** |
|  |  | White (reference) | 5789 (83.64) | 340 (5.9) | 1 |  |
|  | Sex | Female | 3029 (43.77) | 210 (6.9) | 1.149 (0.869, 1.518) | 0.33 |
|  |  | Male (reference) | 3892 (56.23) | 238 (6.1) | 1 |  |

CI, confidence interval
Adjusted OR, adjusted odds ratio: generalized estimating equations were fit, with an “independence” correlation structure, estimating the outcome in the leftmost column, adjusted for age, sex, SOFA first 24h, and source database. OR > 1 means that the racial and ethnic group is more likely to have the outcome, compared to White patients
Bold p-values are significant at $p < 0.05$
Hidden Hyper/Hypothermia (HH)H, either hidden hyperthermia or hidden hypothermia (OR operator)
Hidden Hyperthermia, Oral/Core temperature measurement > 38°C, but temporal measurement ≤ 38°C
Hidden Hypothermia, Oral/Core temperature measurement < 36°C, but temporal measurement ≥ 36°C
The outcomes are defined as 1 if the aforementioned phenomena occur, and 0 otherwise.

**Table 3.** Cohort characteristics (for secondary outcome), grouped by HHH, per pair.

|  | Grouped by Hyper/Hypothermia |  |  |  |
| --- | --- | --- | --- | --- |
|  | Missing | Overall | Hidden | Observed |
| N (%) |  | 434 | 236 (54.4) | 198 (45.6) |
| Time between paired measurements (min), median [Q1,Q3] | 0 | 53.0 [30.0,60.0] | 55.0 [45.0,60.0] | 39.5 [15.0,60.0] |
| Time to pair captured in the ICU, since ICU admission, if in the ICU median [Q1,Q3] | 264 | 47.3 [9.3,131.9] | 64.4 [27.9,170.5] | 29.7 [4.1,107.9] |
| Time to ICU admission, since Hospital admission, median [Q1,Q3] | 264 | 2.1 [0.5,66.7] | 2.9 [1.1,105.9] | 1.8 [0.1,45.9] |
| Temperature, at Oral/Core (°C), median [Q1,Q3] | 0 | 35.7 [34.9,35.9] | 35.8 [35.6,35.9] | 35.3 [34.4,35.9] |
| Temperature, at Temporal (°C), median [Q1,Q3] | 0 | 36.2 [35.8,36.7] | 36.4 [36.2,36.8] | 35.7 [35.3,35.9] |
| SOFA, at pair (previous 24h), median [Q1,Q3] | 269 | 6.0 [4.0,10.0] | 6.0 [3.8,10.0] | 6.0 [4.0,9.0] |
| SOFA, after pair (future 24h), median [Q1,Q3] | 286 | 7.0 [4.0,10.0] | 6.0 [3.0,9.0] | 7.0 [5.0,11.0] |
| Time to ICU discharge (h), median [Q1,Q3] | 0 | 51.2 [19.7,180.5] | 22.1 [18.0,87.4] | 110.4 [41.5,232.9] |
| Time to Antibiotic, after pair (h), median [Q1,Q3] | 127 | 5.1 [2.2,12.6] | 5.0 [2.2,7.2] | 5.7 [2.3,19.0] |
| Time to Culture, after pair (h), median [Q1,Q3] | 256 | 12.5 [3.1,50.1] | 13.0 [4.7,94.9] | 11.4 [2.8,44.9] |
| Time to Lactate, after pair (h), median [Q1,Q3] | 267 | 3.6 [1.5,9.1] | 6.1 [2.5,26.2] | 2.8 [1.2,7.4] |
| Lactate (if present after the pair), median [Q1,Q3] | 267 | 1.8 [1.3,2.9] | 1.7 [1.3,2.1] | 1.9 [1.3,3.7] |
| Temperature measurement captured in the ICU, n (%) | 0 | 172 (39.6) | 58 (24.6) | 114 (57.6) |
| **Oral/Core Temperature Measurement Location, n (%)** |  |  |  |  |
| Bladder | 0 | 4 (0.9) | 2 (0.8) | 2 (1.0) |
| Esophageal |  | 58 (13.4) | 8 (3.4) | 50 (25.3) |
| Oral |  | 305 (70.3) | 207 (87.7) | 98 (49.5) |
| Rectal |  | 67 (15.4) | 19 (8.1) | 48 (24.2) |
| Culture after pair, before discharge, n (%) | 0 | 110 (25.3) | 38 (16.1) | 72 (36.4) |
| Antibiotics after pair, before discharge, n (%) | 0 | 258 (59.4) | 145 (61.4) | 113 (57.1) |
| Lactate after pair, before discharge, n (%) | 0 | 142 (32.7) | 35 (14.8) | 107 (54.0) |
| Culture 48h before pair, n (%) | 0 | 171 (39.4) | 66 (28.0) | 105 (53.0) |
| Antibiotics 48h before pair, n (%) | 0 | 208 (47.9) | 84 (35.6) | 124 (62.6) |
| Lactate 48h before pair, n (%) | 0 | 172 (39.6) | 56 (23.7) | 116 (58.6) |
| Culture Positivity (if present after the pair), n (%) | 324 | 25 (22.7) | 9 (23.7) | 16 (22.2) |

Summary characteristics of the second cohort, stratified by HHH or OHH, to study the secondary outcome: a composite of culture order, antibiotics administration, or lactate measurement, whichever comes first, as a representation of SEP-1 initiation.

### Secondary outcome: Time to initiation of SEP-1 bundle interventions

Compared to patients with OHH, patients with HHH had delayed secondary outcomes, consisting of a composite of culture orders, antibiotics administration, and lactate measurements, whichever happened first. This outcome should flag the initiation of SEP-1 actions as a bundle. Unadjusted cumulative incidence curves can be found in Figure 2. The p-value obtained by the log-rank test was 0.002; the null hypothesis that the time to this secondary outcome was the same across OHH and HHH patients is thus rejected. Finally, there were no significant differences in culture positivity [positive cultures: 9 (23.7%) versus 16 (22.2%), *p*=1] between HHH and *OHH* patients, respectively. (Table 3)

## Discussion

In our large, multicenter retrospective study involving a large and diverse patient population (n = 4,144), we investigated racial disparities in temperature measurement using the temporal method versus oral/core measurements. Our findings revealed that Asian, Black, and Hispanic populations are more prone to *Hidden Hyper/Hypothermia* identification when using temporal thermometers compared to White patients. This delay was associated with significantly lower compliance with sepsis management guidelines (SEP-1), as there were delays in obtaining blood cultures, antibiotic ordering, or lactate levels. To our knowledge, this is the first study to associate temporal thermometers with racial and ethnic disparities to guidelines compliance, highlighting the urgent need to address these disparities to improve patient care and to better adhere to sepsis management protocols.

Our study builds on the work of Bhavani et al.^11^. These investigators conducted a retrospective analysis in four hospitals within the Emory health system, including 4,375 patients identified as either White or Black, and excluded patients who identified as Asian and Hispanic/Latinodue to small sample sizes. In contrast, our study was able to include these racial and ethnic groups because of its much larger and nationwide sample. While Bhavani and colleagues focused on patients with suspected infections, our study covered all hospitalized patients, evaluating the impact of racial and ethnic disparities on adherence to SEP-1 guidelines. We also included hypothermia, which is equally important in sepsis identification and management.^21^ While we confirm Bhavani et al.’s^11^ findings of temporal thermometry disparities in Black patients, we also discovered that these disparities extend to other minority groups and include hypothermia. As we link thermometry inaccuracies to treatment initiation delays, our study suggests that solely relying on temporal thermometers can lead to racial disparities and delay treatment onset, hampering compliance with SEP-1 and reimbursement.

Past work demonstrated that racially minoritized patients experience a higher incidence of sepsis and elevated mortality rates compared to White patients.^22^ This disparity has been attributed to clinical characteristics, variable quality of hospitals, genomic differences, socioeconomic status, and environmental factors.^16,23^ Our study adds a novel perspective by examining device biases as a potential contributor in sepsis disparities which is seldom reported in literature.^24^ Similarly to pulse oximeters, temporal thermometers use infrared light, which might explain the disparities observed in both Bhavani et al.’s^11^ study and ours. Differences in skin tone are suggested to be the reason for the observed disparities, as these devices have been primarily validated on lighter-skinned populations, potentially leading to less accurate readings for individuals with darker skin tones.^12,13^ Similar to our findings of delayed treatment and potentially delayed diagnosis due to temporal thermometer disparities, disparities in pulse oximetry have been found to correlate with delayed diagnosis of severe pneumonia, delayed admission, and treatment initiation.^13,25^

With the new CMS rules starting in 2024, including SEP-1 compliance under the VBP program for reimbursement, racial disparities in the accuracy of temporal thermometers have significant patient and hospital consequences.^7^ CMS utilizes an electronic algorithm to retrospectively extract the earliest time the sepsis criteria were met followed by initiation of the SEP-1 bundle.^26,27^ Our results indicate that discrepancies in temporal readings, which can mean missed SIRS cut-off temperature measurements, were associated with delays in the initiation of SEP-1 actions. Although those patients had oral/core temperature readings exceeding the SIRS cut-off, there were still significant delays in initiating the SEP-1 bundle. From a patient perspective, delayed identification of sepsis and initiation of the SEP-1 bundle leads to poorer outcomes, including increased resource utilization, longer lengths of stay, and higher mortality rates, all of which negatively impact patient health and hospital economics.^28–30^ Furthermore, clinical teams treating patients with HHH may be unable to claim appropriate billing for sepsis, further complicating compliance and reimbursement issues. Delaying treatment in septic shock increases the risk of complications, leading to higher resource utilization and negatively impacting payment structures. ^3132^

Until definitive solutions by improved devices are developed, considering the available evidence, clinicians should consider incorporating a 0.1–0.3°C difference towards critical thresholds in temporal readings in minority groups compared to oral/core temperature measurements.^33–36^ If sepsis is under suspicion, it is reasonable to prioritize oral or core temperature measurements over temporal readings whenever possible or to implement frequent validation of temporal thermometers against oral/core measurements to ensure accuracy and minimize the risk of delayed recognition. Future studies should examine further clinical situations, such as among mechanically ventilated patients, as well as the variability in differences between temporal and oral/core readings to determine if these differences remain stable throughout the entire admission, which could help in developing correction models. Incorporating quantitative measurements for skin tone and pigmentation into such modeling could also be promising, similar to work that has been initiated in pulse oximetry. ^37,38^

This study has several limitations. Because it is retrospective, pairing temperature measurements can introduce selection bias, as measurements depend on clinical suspicion, hospital practices, and a patient’s illness severity — highlighting the need for prospective studies to systematically address such disparities. Our exploratory approach underscores the importance of this issue and encourages further investigation. We also excluded fluid balance data and antipyretic medications due to extraction challenges or database unavailability, potentially limiting the full characterization of patient clinical status. Although we included all hospitalized patients, excluding those without a recorded SOFA score skewed our dataset toward ICU patients, which may reduce generalizability to non-ICU settings. Additionally, limited sample sizes of Hispanic and Asian patients constrained our ability to draw robust inferences for these groups, mirroring the limitations of the prior Emory cohort. Their inclusion, however, emphasizes the need for larger, more representative samples to address these gaps. Lastly, future studies should validate our findings in other health systems and explore how temporal thermometer inaccuracies correlate with clinical outcomes such as organ dysfunction, length of stay, or mortality.

## Conclusion

Our study highlights significant racial and ethnic differences in temperature measurement obtained by the increasingly popular temporal thermometer method. These disparities have implications in the adherence to sepsis management protocols, with consequences in exacerbating existing health disparities and in hospitals’ reimbursement programs, like the CMS SEP-1 bundle. Validating temporal temperature readings with gold standard measurements may be an important strategy to address these disparities, improve patient care, and optimize health systems.

## Data Availability

MIMIC and eICU data are publicly available on PhysioNet.

https://mimic.mit.edu/

## Supplemental Text

MIMIC-IV and eICU-CRD are publicly available on PhysioNet.

### Duke Health

Patients admitted to an adult intensive care unit (ICU) at Duke University Health System, a teaching center with more than 1,500 beds in total, located in the South of the United States. Data was extracted from Epic through the Protected Analytics Computing Environment (PACE), including 2014 – 2024.

### MIMIC-IV

MIMIC-IV extends MIMIC-III to include patients admitted to the ICU from 2008 to 2022. Approximately 80,000 de-identified medical records are archived in the MIMIC-IV database. ^41^

### eICU-CRD-1

The eICU-CRD database is a multi-center database sourced from the Philips Healthcare eICU Telehealth Program. It contains information about over 200,000 admissions from 208 hospitals or ICUs monitored by eICU programs across the United States, between 2014 and 2015. There is no overlap with MIMIC. ^42^

### eICU-CRD-2

eICU-CRD-2 extends eICU-CRD-2 to include patients from 2019-2022, i.e, including COVID-19 years.

### Antibiotics

Antibiotics were defined as antibiotics meeting Adult Sepsis Event criteria by the Hospital Toolkit. ^43^ These were matched by both medication name and route and reviewed by a clinician. Medication rows that did not have both a medication name and route were ignored.

## Supplemental Figures

**Supplemental eFigure 1.**
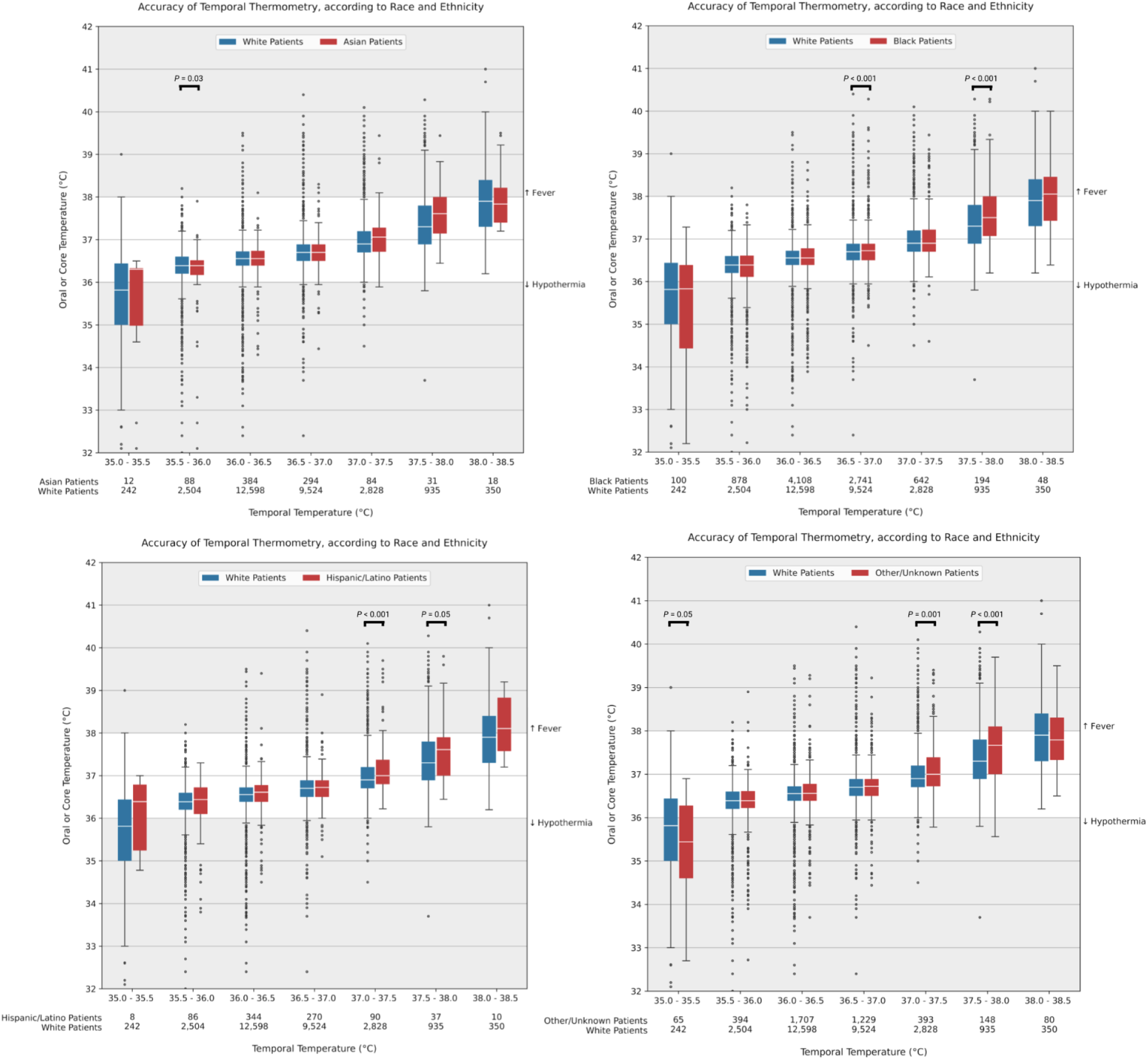
Accuracy of Temporal Thermometry, compared to Oral/Core Thermometry, according to race and ethnicity, when using all temperature pairs (before exclusion for primary or secondary outcome) Based on Sjoding et al. ^40^ 2-sided t-tests between the distributions of Asian, Black, Hispanic/Latino, and Other/Unknown versus White patients’ contact temperature in each 0.5°C bin of temporal temperature were performed. Significant p-values are highlighted.

**Supplemental eFigure 2.**
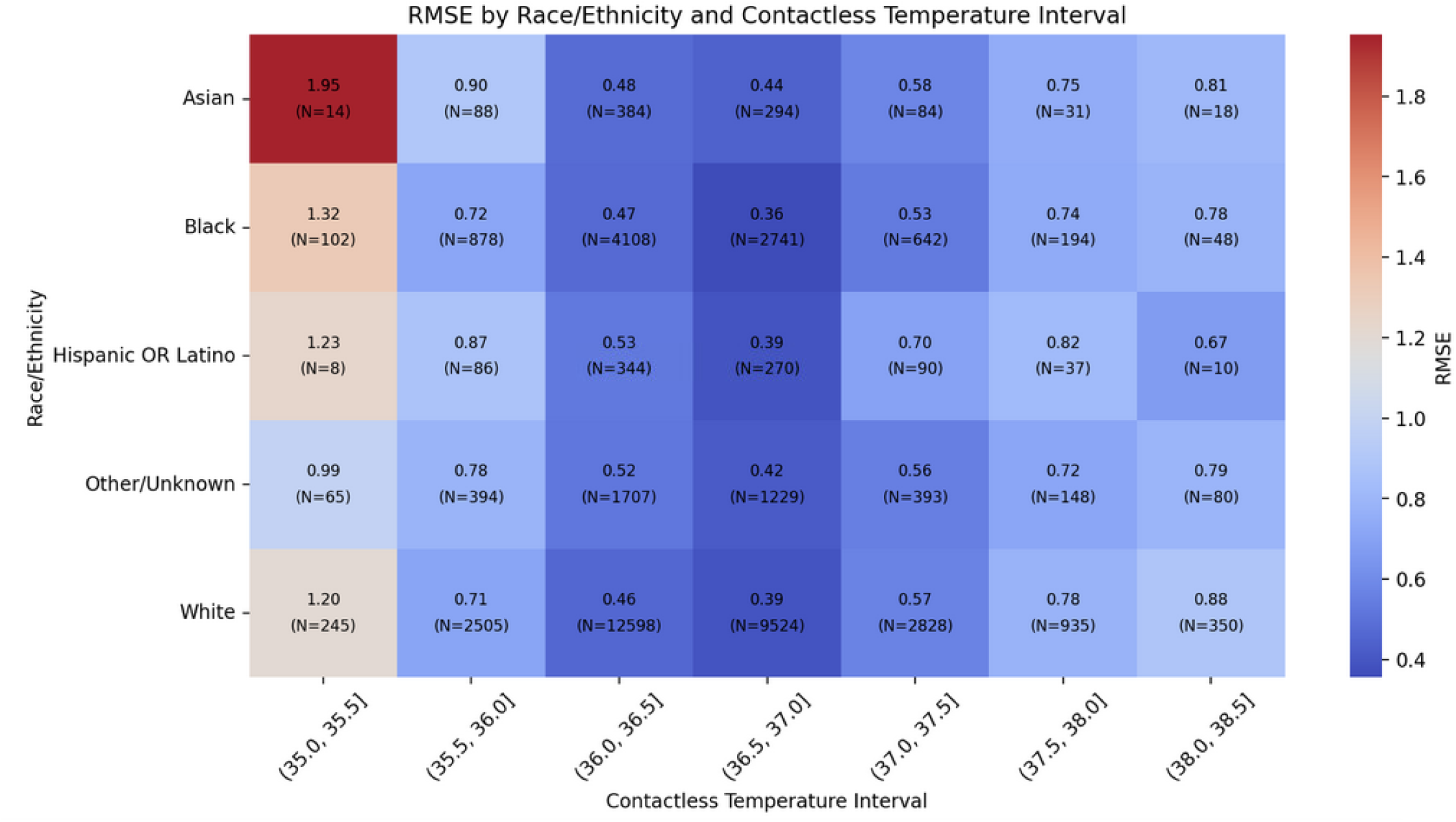
Root mean square error of temporal temperature when compared to oral/core, per race and ethnicity and temperature range, when using all temperature pairs (before exclusion for primary or secondary outcome)

**Supplemental eFigure 3.**
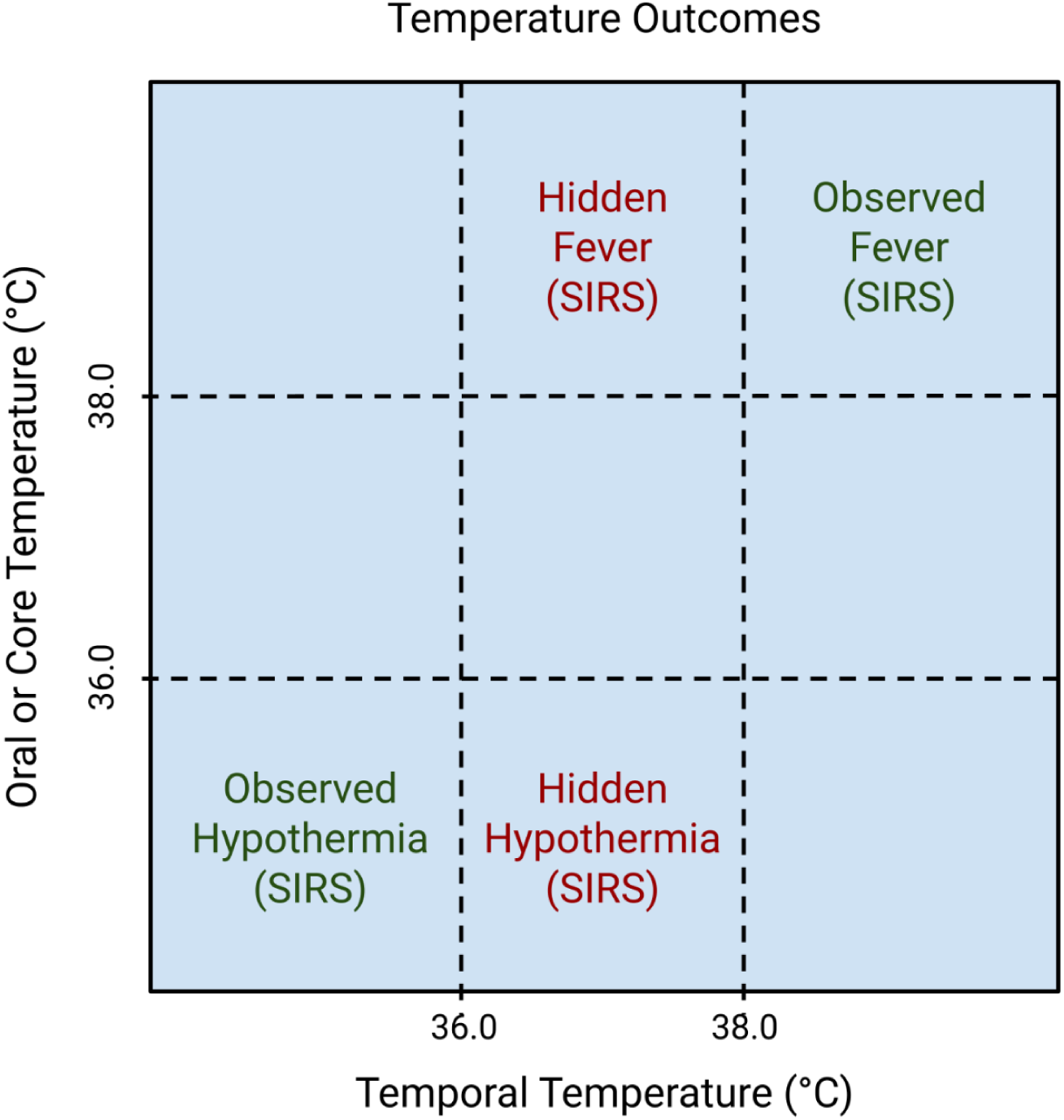
Temperature Outcomes

